# Guillain-Barr é syndrome in COVID-19: A scoping review

**DOI:** 10.1101/2020.06.13.20130062

**Authors:** Imran Ahmad, Farooq Azam Rathore

## Abstract

**Introduction:** The novel corona virus (COVID19) can result in several neurological complications. Guillain-Barré Syndrome (GBS) is one of them and has been reported from different parts of the world in this pandemic. It is an acute post infectious polyneuropathy. The review aims to summarize the demographic features, clinical presentation, diagnostics workup, and management strategies of COVID-19 associated GBS reported in literature.

**Material and method:** We searched Medline, PubMed Central, SCOPUS and Google Scholar using pre-defined keywords, with no time limits and in English language only. We aimed to include all kind of manuscripts. Last search was done on 18^th^ May 2020.

Demographics, clinical features, diagnostic workup, management, and outcomes were documented in the data sheet.

**Results:** We identified 24 cases of COVID-19 associated GBS. Most of the cases were reported from Italy followed by USA. Majority were males (18 /24) The age ranged from 23 -84 years. The clinical presentation was typical sensory-motor GBS in most. Nine patients had facial palsy of which five had bilateral involvement. Two patients had bilateral abducent nerve palsy while two presented as paraparetic GBS variant with autonomic dysfunction. Electrodiagnostics was performed in 17 patients only and 12 had typical features of acute inflammatory demyelinating polyneuropathy.. Intravenous immunoglobulins were the preferred mode of treatment in most of the patient. There was one death, and most were discharged to rehabilitation or home.

**Conclusion:** GBS is a frequent neurological complication associated with COVID-19. There is no clear causative relationship between GBS, and COVID-19 at present and more data are needed to establish the casualty. However, most cases have a post-infectious onset with male preponderance. Most of the cases have a typical presentation but some may present in an atypical way. Prognosis is generally good.

## Introduction

The novel coronavirus (COVID19) infection originated from Huanan seafood market in Wuhan city China in December 2019. It rapidly spread to more than 200 countries of the world with 7 million confirmed cases and more than 400,000 fatalities as of 10^st^ June 2020.^1^ COVID-19 primarily affects the respiratory tract and the lungs. However, involvement of cardiovascular, renal system and neurological system has also been reported. The reported neurological manifestations and complications of COVID-19 include anosmia, headaches, dizziness, delirium, stroke, epilepsy, encephalitis, encephalopathy, myalgias and Guillain-Barr é syndrome (GBS) ^2,3,4^ At present, there are no reviews, full length research article or reports discussing a specific neurological complications in detail.

The objective of this review is to summarize the important demographic features, clinical presentation, diagnostics, and management strategies of COVID-19 associated GBS reported in literature so far. We aim to inform the readers about this important neurological manifestation of COVID-19 in order to formulate better diagnostic and management strategies.

### Historical Perspectives on Guillain-Barr é syndrome

The mysterious link between infections followed by paralysis has intrigued physicians for centuries.^5^ Although this relationship was mentioned by the famous Muslim physician Avicenna centuries ago and many other authors published their cases too, but the detailed description of the disease, including nerve conductions studies and cerebrospinal fluid analysis with albuminocytological disproportions was first documented by three French physicians Georges Guillain, Jean Alexandre Barr’, and Andr’e Strohl, who were working together at the Neurological Center of the French 6th Army during first world war. They treated and published data on two soldiers, who developed typical features of GBS with complete recovery. The disease was named Guillain-Barr é syndrome after them. ^6^

### Pathophysiology and Clinical Features of Guillain-Barr é syndrome

GBS is acute onset immune mediated disorder characterized by rapidly progressive limbs and bulbar weakness which can lead to respiratory failure. ^7^ Many triggers for GBS have been identified including bacterial and viral infections, surgery, and pregnancy. The link of GBS with vaccination is controversial. Respiratory and Gastrointestinal infections constitute two third of cases. The molecular mimicry between the cell membrane antigen of microbe and ganglioside component of nerve antigen misdirects the immune response. This immune response is humoral mediated and not T cell mediated. The prototype example is of Campylobacter Jejuni infection. The carbohydrate moiety of liopooligosacchrides of Campylobacter Jejuni is capable of inducing antibodies that cross react with glycans present on nerve gangliosides.^8^ The exact trigger to mount this misdirected immune response is still not known. There is no specific genetic predisposition as only 1% of all campylobacter infections will result in GBS. GBS has also been reported after viral infections for example Cytomegalovirus, Ebstein-Barr virus, Influenza, Zika, and Chikungunya virus.12^,8^

The clinical hallmark is hyporeflexia or areflexia. The course of the disease is monophasic. Recommended treatment for GBS includes plasmapheresis (PLEX) and immunoglobulins (IVIG) infusion.^9^ GBS was initially described only as a demyelinating polyneuropathy. Typical features of demyelination on electrodiagnostic studies (EDX) include prolong distal latencies, reduction of conduction velocity, prolong F-waves, temporal dispersion, and conduction block. Clinically many different variants with distinct clinical and electrophysiological features have been reported in literature^2,10^. These include, cranial, autonomic, ataxic, paraparetic and mixed variety. The GBS reported from North America and Europe is predominantly demyelinating type while in Asian countries the axonal type of GBS constitutes 30-50 % of the reported cases. ^11,12,13, 14^ The mortality of GBS reported from European and North American studies ranges from 3-7%, and is mainly due to respiratory failure, deep vein thrombosis and autonomic dysfunction. The axonal variants of GBS mostly reported in the Chinese and Asians populations have a poor prognosis. ^15^ The poor prognosis is because peripheral nerve remyelination is a natural and active process. Whereas in case of axonal variant of GBS, once the axonal integrity is damaged, it does not be regenerate actively and completely.^2^

### Basic Virology of SARSCoV-2

SARS-CoV-2 belongs to the broad family of Coronaviruses sub-family beta Coronaviruses ^16^. These are enveloped, single-stranded RNA viruses with a diameter of approximately 60–140 nm. The viral envelope has four structural proteins known as S (spike), E (envelope), M (membrane), and N (nucleocapsid). It is the interaction with the spike protein and host cell receptor which is essential for virulence and infectivity.^17^

### Neurological injury due to Coronavirus

Coronaviruses are not primarily neurotropic viruses and their primary target is respiratory and cardiovascular systems. It is through the Angiotensin-converting enzyme 2 (ACE-2) receptors the virus is attached to host cells leading to internalization and subsequent viral replication. This receptor is also found in glial cells in the Central Nervous System (CNS) and spinal neurons. Very rarely the virus can invade peripheral nerves and lead to retrograde transfer via synapse mediated route to CNS. Another proposed route of entry is through the olfactory nerves. ^18, 19^ Past experience with Severe Acute Respiratory Syndrome (SARS) and Middle East Respiratory syndrome (MERS) related cases has also provided insights into the neuro invasive potential of Coronaviruses.^20, 21^ As the number of COVID-19 cases with neurological manifestations and complications are being reported more frequently, there is growing evidence for neurotoxic potential of COVID-19. This neurotoxicity can occur as a result of direct or indirect insult by virus and may manifest in form of post-infectious complications like GBS. In this review we will focus only on the post infectious complication only.

### Mechanism of GBS in coronavirus infection

COVID-19 does not directly invade peripheral nerves, nerve roots, or anterior horn cells leading to inflammation and death of motor neurons as seen in polio virus or West Nile virus. Even the Cerebrospinal fluid (CSF) Polymerase chain reaction (PCR) for coronavirus in multiple reported cases of COVID-19 related GBS has been negative.^22^ It is likely a post infectious or may be a para-infectious complication resulting from an aberrant immune response. During the inflammatory phase numerous mediators of inflammation are released from activated leukocytes including Interleukin-6 (IL-6), named as Cytokine storm. This can result in major organ damage, rapid deterioration of the patient and ultimately death.^23^ However, due to lack of experimental data It is difficult to deduct if IL-6 is also responsible for the c is also causing neurological damage ^37^ After the acute phase of the infection, an immune response is generated by the host and may lead to a misdirected response against host epitopes. It can result in an autoimmune, response directed against peripheral nerves and nerve roots in susceptible individuals. This may be either demyelinating or axonal degeneration type. This results in a typical GBS like presentation in the peripheral nerves and spinal roots. However, due to lack of clear data, there is still not enough evidence available to conclude if antibodies to any specific ganglioside antigen are present in these cases or not.

### Literature Search strategy

We searched Medline, PubMed Central, SCOPUS and Google Scholar using keywords “COVID-19”, “Coronavirus”, “Coronavirus Infections”, “Coronaviridae”, “2019 nCoV “. “pandemic”, “SARS-COV-2”, “neurology”, “neurological”, “complications”, “manifestations”, “Guillain-Barr é syndrome”, “GBS”, “acute inflammatory demyelinating polyneuropathy”,” Demyelinating Polyradiculoneuropathy”, “polyneuropathy”, and “Miller Fisher syndrome”. Different combinations of Boolean logic (AND, OR and NOT) were used to identify relevant articles. Search was limited only to English language manuscripts with no time limit. It is important to note that new data is being shared regularly and so far, it consists mostly of pre-prints, letters to editor, single case reports, small case series, and part of an article describing clinical features of COVID-19. Most of the data on COVID-19 at present is published from countries most severely affected, including China, Italy, Spain, and USA. The last literature search was done on 18^th^ May 2020. At that time there was no specific research article, systematic or narrative review describing COVID-19 associated GBS. However, we identified two systematic reviews protocols on this topic registered in the International prospective register of systematic reviews. ^24, 25^

Both authors independently performed the literature search and compared the results for any major discrepancies. The information was extracted on a pre-designed data sheet. The items of interest were the demographic data, presenting features, clinical examination, laboratory and radiological investigations, treatment protocol and outcomes. Due to the limited number of cases and nature of the review, a quantitative analysis was not done, and we have only provided a qualitative review of the retrieved information.

## Results

### Characteristics of included studies

After removing the duplicates, non-English manuscripts, and unrelated articles, we identified 24 cases of GBS in COVID-19, published in the English biomedical literature till 18^th^ May 2020. These were published as letter to editor, case reports or case series. The results are summarized in the Tables 1 and 2.

**Table 1:** Details of the Demographics and clinical features of COVID-19 related GBS.

|  | Country | Author | Age<br>(Years) | Gender | Presenting symptoms | Clinical Examination | H/o Respiratory or GIT<br>infection | Travel history |
| --- | --- | --- | --- | --- | --- | --- | --- | --- |
| 1. | China | Zhao | 61 | Female | Acute weakness both legs,<br>Severe fatigue | Symmetrical weakness grade 4/5 on<br>MRC Scale<br>Areflexia<br>After three days power 3/5 legs<br>Sensations decreased to light touch in<br>feet | Develop fever and cough on<br>eight day of illness. | Travel to Wuhan City, China |
| 2. | Iran | Sedaghat | 65 | Male | Started from lower limbs<br>Five days upper limbs involved<br>symmetrical ascending<br>quadriparesis,<br>Bilateral facial palsy | UL Muscle Power 2/5 proximal and<br>3/5 distal<br>LL Muscle Power 1/5 proximal and 2/5<br>distal<br>Areflexia<br>Impaired vibration and proprioception<br>distally (DM) | Cough, Fever, and dyspnea<br>02 weeks prior to admission | Not Reported |
| 3. | France | Camdessa<br>nche | 65 | Male | paresthesia hands and feet<br>progressed to Quadriplegia in<br>3 days | 2/5 in legs and arms proximally. 3/5<br>forearm and 4/5 hand<br>Areflexia<br>Absent vibrations<br>Dysphagia present | Cough and fever 11 days<br>before admission | No |
| 4. | USA | Virani | 54 | Male | Numbness and weakness. | Power 4/5 LL initially | Fever at presentation. | No |
|  |  |  |  |  | followed by Urinary retention and ascending paralysis | Progressed to 2/5 Areflexia | Cough for 10 days |  |
| 5. | Spain case 1 | Gutiérrez | 50 | Male | Vertical diplopia, Gait instability, perioral paresthesia Headache | Ataxia and Areflexia<br>Right INO, Right 3<sup>rd</sup> nerve palsy<br>No facial weakness<br>Anosmia and ageusia | Five days back<br>Fever, cough<br>Malaise and backache | No |
| 6. | Spain case 2 | Gutiérrez | 39 | Male | Diplopia and ageusia | Bilateral abducent palsy<br>Areflexia<br>No ataxia and Gait instability | Fever and diarrhea 3 days before admission. No respiratory symptoms<br>No respiratory symptoms | No |
| 7. | Italy | Toscano, | 77 | Female | Paresthesia in lower limbs and hands, Acroparesthesia and hypogeusia | Flaccid quadriplegia Areflexia<br>Later on dysphagia and respiratory difficulty | Fever and cough 7 days before admission | No |
| 8. | Italy | Case 2 | 23 | Male | Facial weakness, mastoid pain and lower limb paresthesia | Hypogeusia<br>Areflexia<br>Bilateral facial palsy<br>Sensory ataxia | Fever and sore throat for 10 days before admission | No |
| 9. | Italy | Case 3 | 55 | Male | lower limb weakness, Paresthesia, and neck pain | Areflexia<br>Quadriplegia<br>bilateral facial palsy | Fever and cough for 10 days before admission. | No |
| 10 | Italy | Case 4 | 76 | Male | Back pain, lower limb weakness and anosmia | Areflexia<br>Quadriparesis | Dry cough for five days before admission and weakness on ninth day | No |
| 11 | Italy | Case 5 | 64 | Male | Cough, asthenia, hyposmia and hypogeusia I followed by proximal weakness and paresthesia | Areflexia paraparesis later on Quadriplegia, facial palsy, bulbar weakness and dysphagia | Cough seven days before onset of neurological symptoms | No |
| 12 | Italy | Alberti | 71 | Male | Paresthesia, Distal weakness progressing to quadriparesis in 3 days | Symmetric limb weakness 2/5 LL and 3/5 UL.<br>Glove and stocking paresthesia and Areflexia | Fever for few days in the previous week | Not Reported |
| 13 | Italy | Padroni | 70 | Female | Paresthesia<br>Asthenia and gait difficulty | 4/5 power<br>Areflexia<br>Sensations intact | 24 days back fever and cough | No |
| 14 | USA | Dinkin | 36 | Male | Bilateral distal Leg Paresthesia and diplopia | Left eye ptosis<br>Mydriasis and partial 3<sup>rd</sup> nerve.<br>Bilateral abducent palsy<br>Gait ataxia, hypoesthesia and areflexia | Fever, cough and myalgias four days before admission. | Not Reported |
| 15 | Switzerland | Ceon | 70 | Male | Paraparesis. allodynia, Myalgia, Difficulty in voiding and constipation | Bilateral lower limb Flaccid paraparesis<br>Areflexia in all limbs<br>Planters down going | Myalgia, fatigue, dry cough 10 days before admission | No |
| 16 | Morocco | Otmani | 70 | Female | Tingling and rapidly progressive weakness | Areflexia and quadriplegia | Dry cough and fever three days before admission total 10 days before first symptom | Not Reported |
| 17 | Italy | Ottaviani | 66 | Female | Difficulty walking and fatigue.<br>Rash on hands | Initially symmetric paraplegia<br>UL power 4/5<br>Unilateral facial palsy<br>areflexia | Mild fever and cough 10 days before presentation | Yes |
| 18 | Germany | Pfefferkorn | 51 | Male | Progressive upper and lower limb weakness<br>Acroparesthesia | 2/5 Muscle power with Quadriplegia<br>Areflexia<br>Later, locked in syndrome<br>Bilateral facial Palsy, Bilateral hypoglossal paresis<br>Complete sensory loss | Fever and flu, 02 weeks ago. | Not Reported |
| 19 | Germany | Scheidl | 54 | Female | Paraparesis,<br>Distal numbness and tingling<br>Later on, developed dysphagia | Proximal 3/5<br>Distal 4/5<br>Areflexia | PCR positive after positive contact 3 weeks before. No symptoms of cough and fever<br>However, had anosmia and ageusia | Not Reported |
| 20 | Iran | Ebrahimzadeh- Case 1 | 46 | Male | Pain, and numbness in distal lower and upper extremities for a week followed by ascending paralysis. | Mild facial nerve palsy on the right side. Muscle Power 4/5 LL progressed to 3/5<br>UL power 5/5 progressed to 4/5<br>Areflexia | Sore throat, dry cough, and mild dyspnea 18 days prior to developing neurological symptoms. | Not Reported |
| 21 | Iran | Ebrahimzadeh- Case 2 | 65 | Male | Ascending upper and lower extremity weakness and paresthesia. | Muscle Weakness 2/5 proximal and 3/5 distal UL<br>UL power 4/5<br>Areflexia in lower limbs<br>UL reflexes +1 | Fever and cough ten days before PCR | Not Reported |
| 22 | USA | Chan-Case 1 | 68 | Male | Gait disturbance<br>Paresthesia hands and feet | 4/5 power hip flexors<br>Absent vibratory and proprioceptive sense at the toes and areflexia in LL and present in UL<br>Later, bilateral fascial palsy, dysphagia, dysarthria and neck flexion weakness<br>Dysarthria and Gait problem | Fever and Cough 18 days back | Not Reported |
| 23 | USA | Chan - Case2 | 84 | Male | Paresthesia of hand and feet<br>07 days back<br>03 days gait disturbance | Power 3/5 proximally, unable to walk independently<br>Gradually progressed bilateral facial palsy<br>Autonomic dysfunction, areflexia in LL and present in UL | 23 days before fever and cough | Not Reported |
| 24 | Spain | Caamaño | 61 | Male | Facial Muscles weakness | Bilateral facial nerve palsy with absent blink reflex<br>Rest of the neurological examination including reflexes were normal. | Fever and cough without dyspnea 10 days prior to admission | Not reported |

ADIP: Acute Inflammatory Demyelinating Polyneuropathy; AMSAN: Acute Motor Sensory Axonal Neuropathy; AMAN: Acute Motor Axonal Polyneuropathy; CSF: Cerebrospinal fluid; GIT: Gastrointestinal tract; IVIG: Intravenous Immunoglobulins; HRCT; High Resolution Computed Tomography scan; LL: lower limbs; ;MRC: Medical Research Council; MRI: Magnetic Resonance Imaging; PCR: Polymerase Chain Reaction; PLEX: Plasma Exchange; UL; upper Limb

**Table 2:**
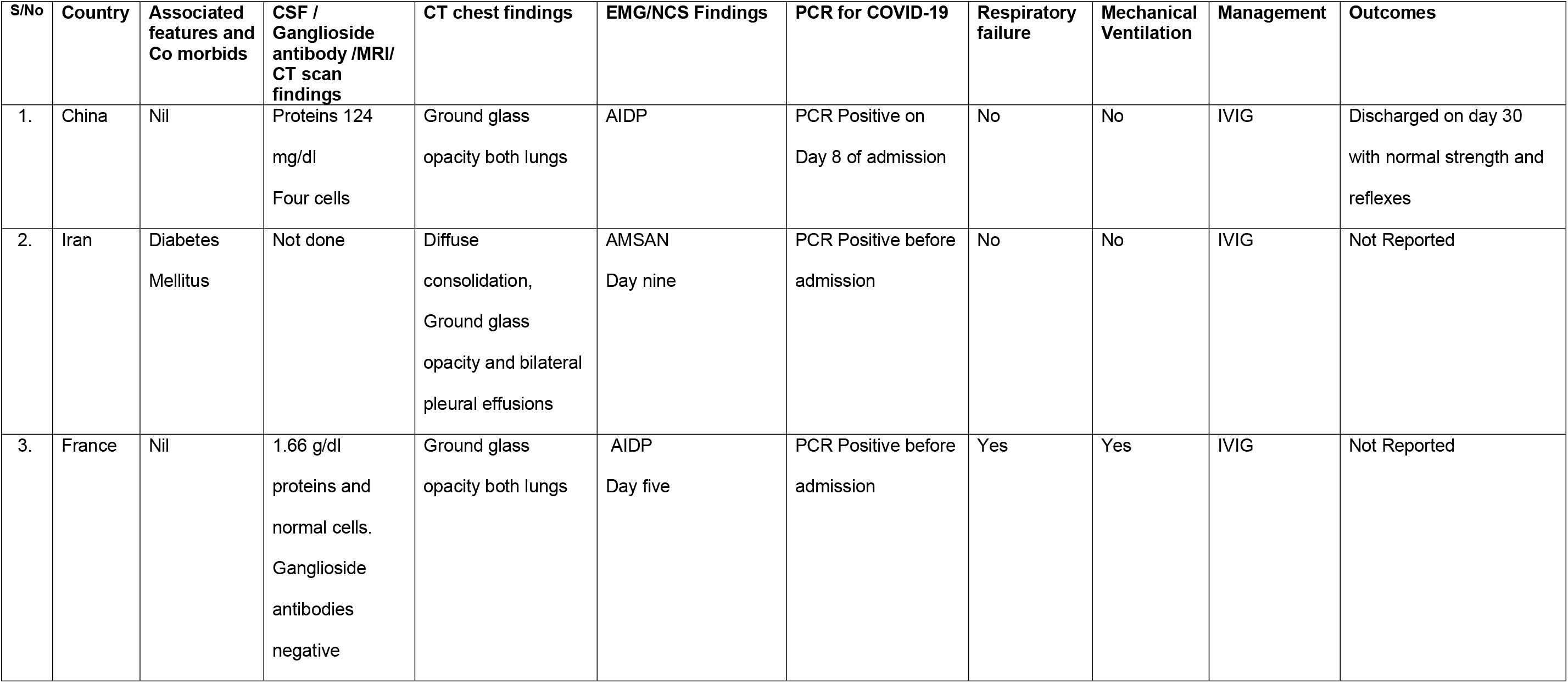
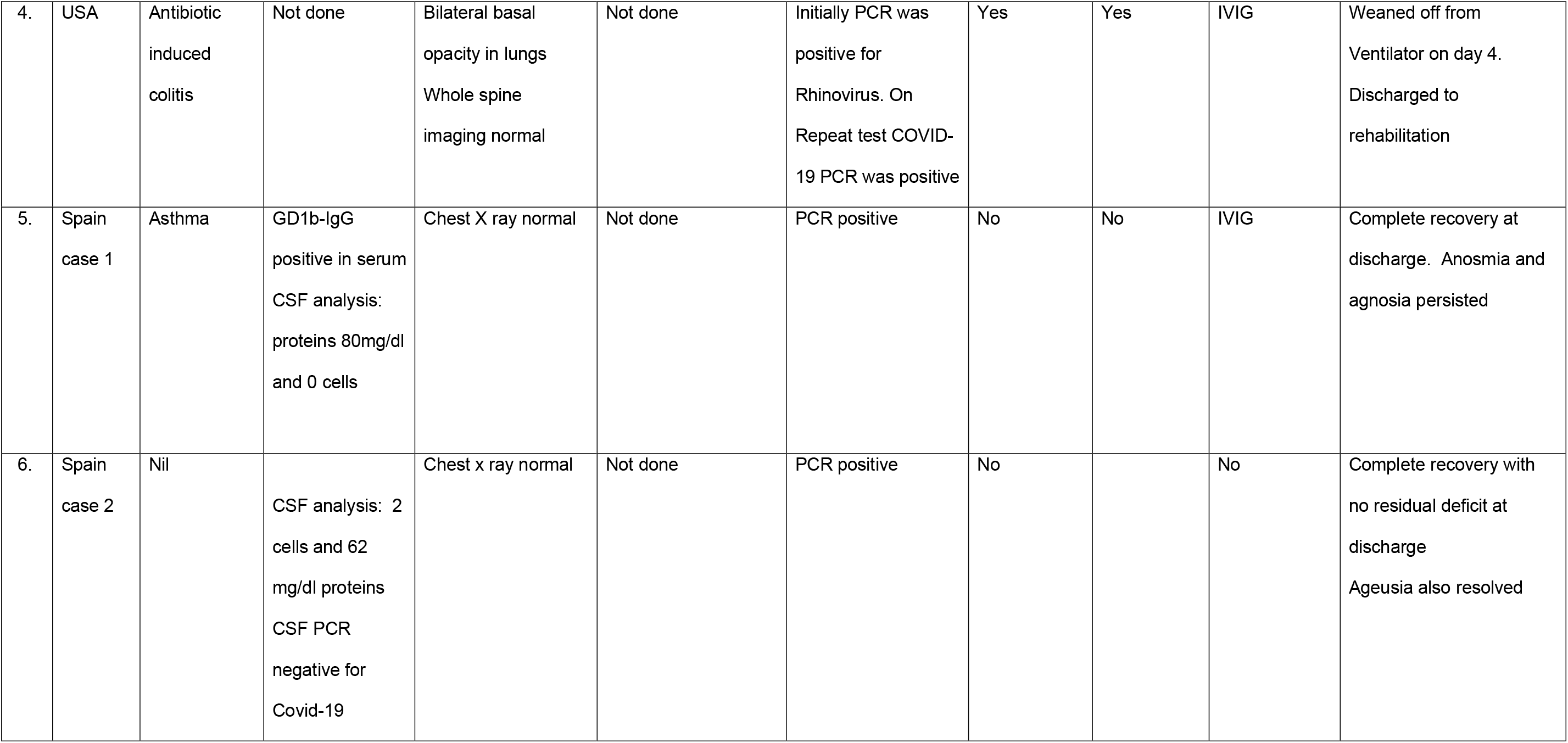
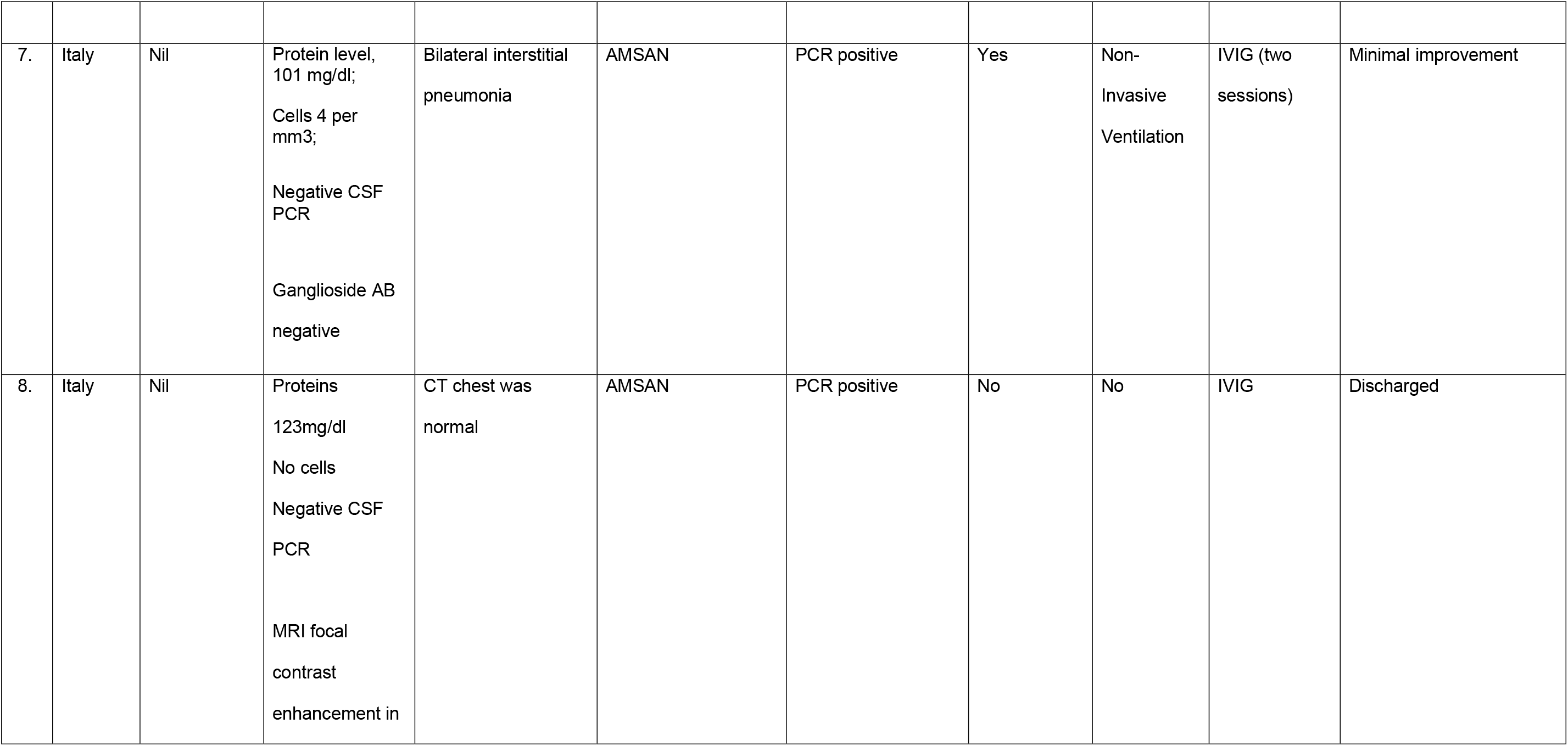
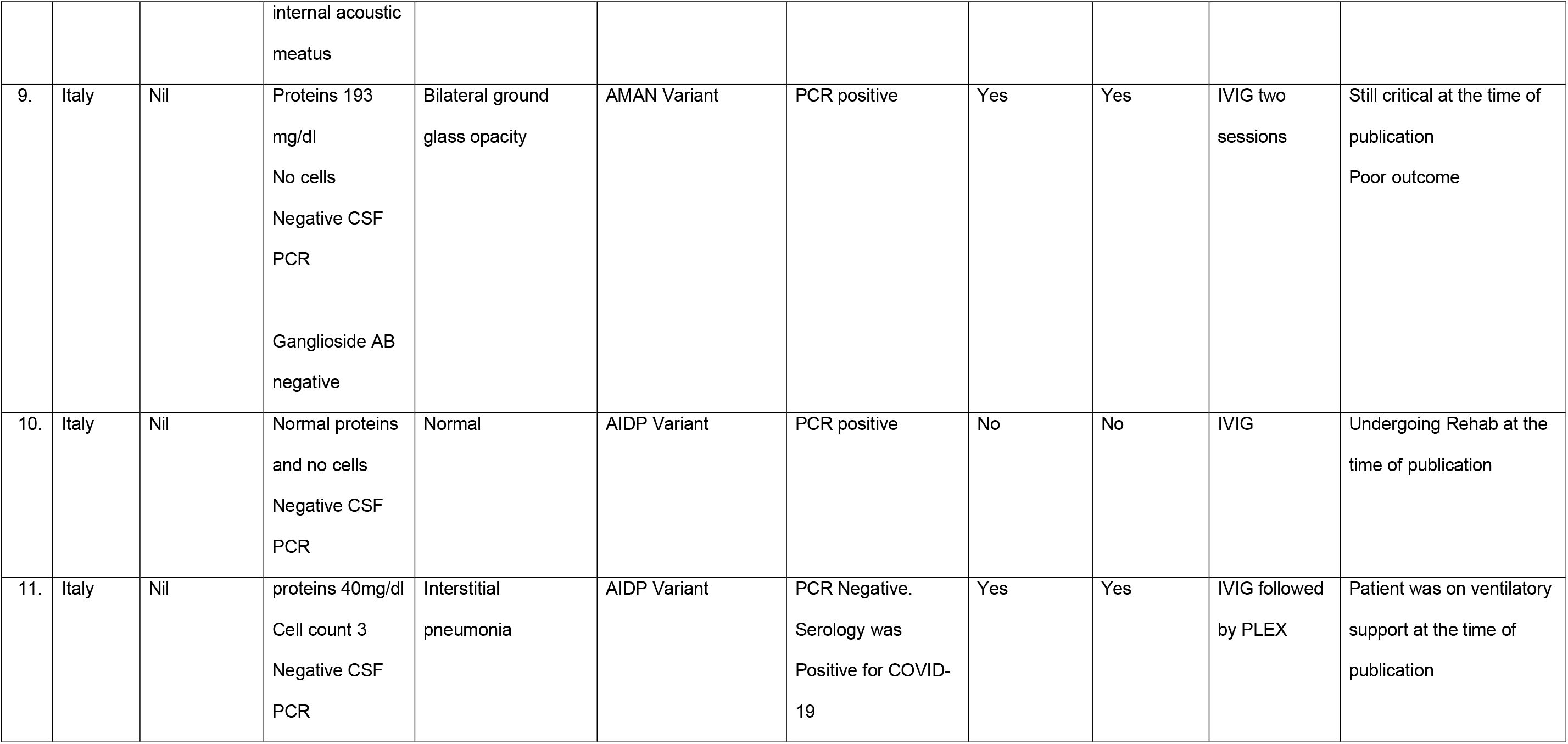
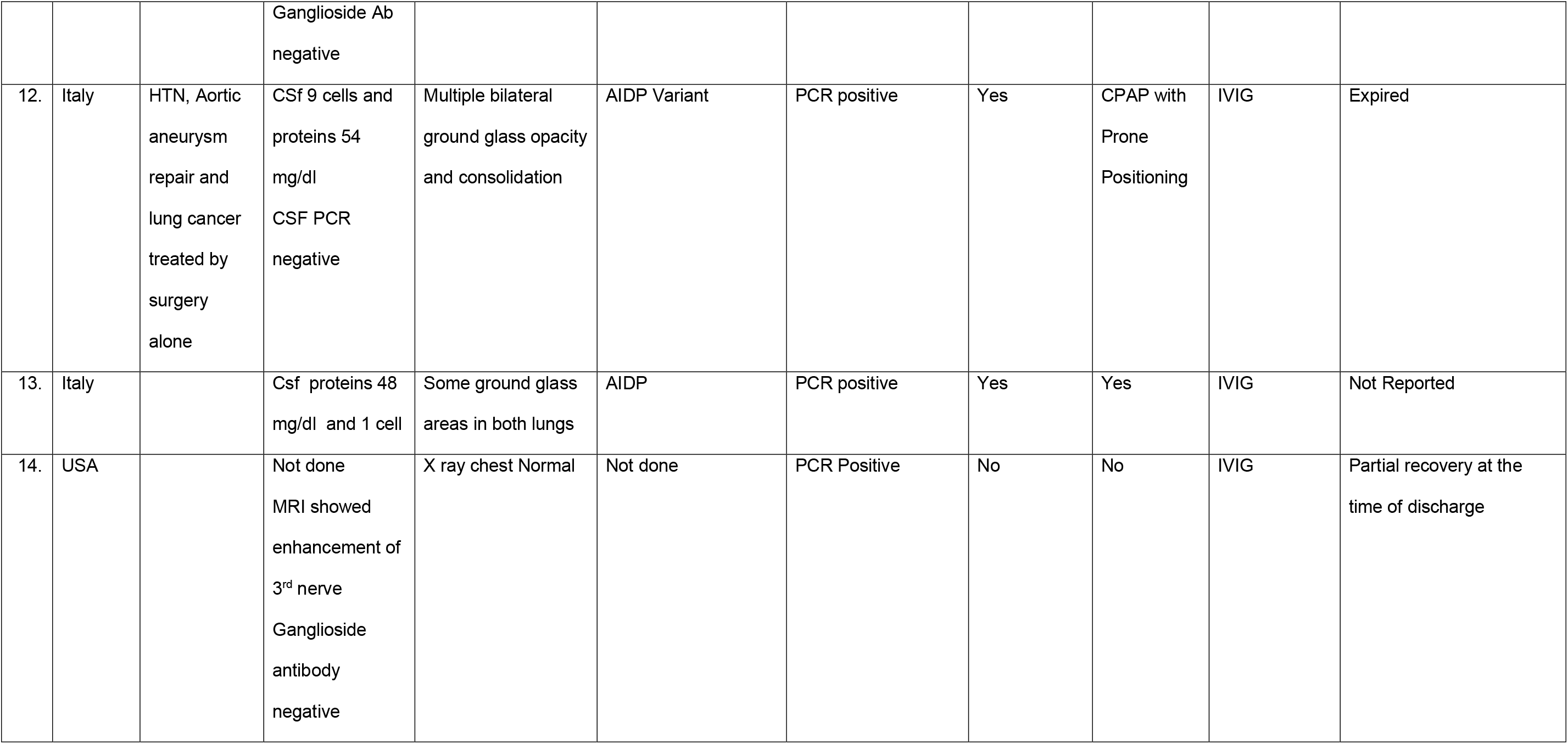
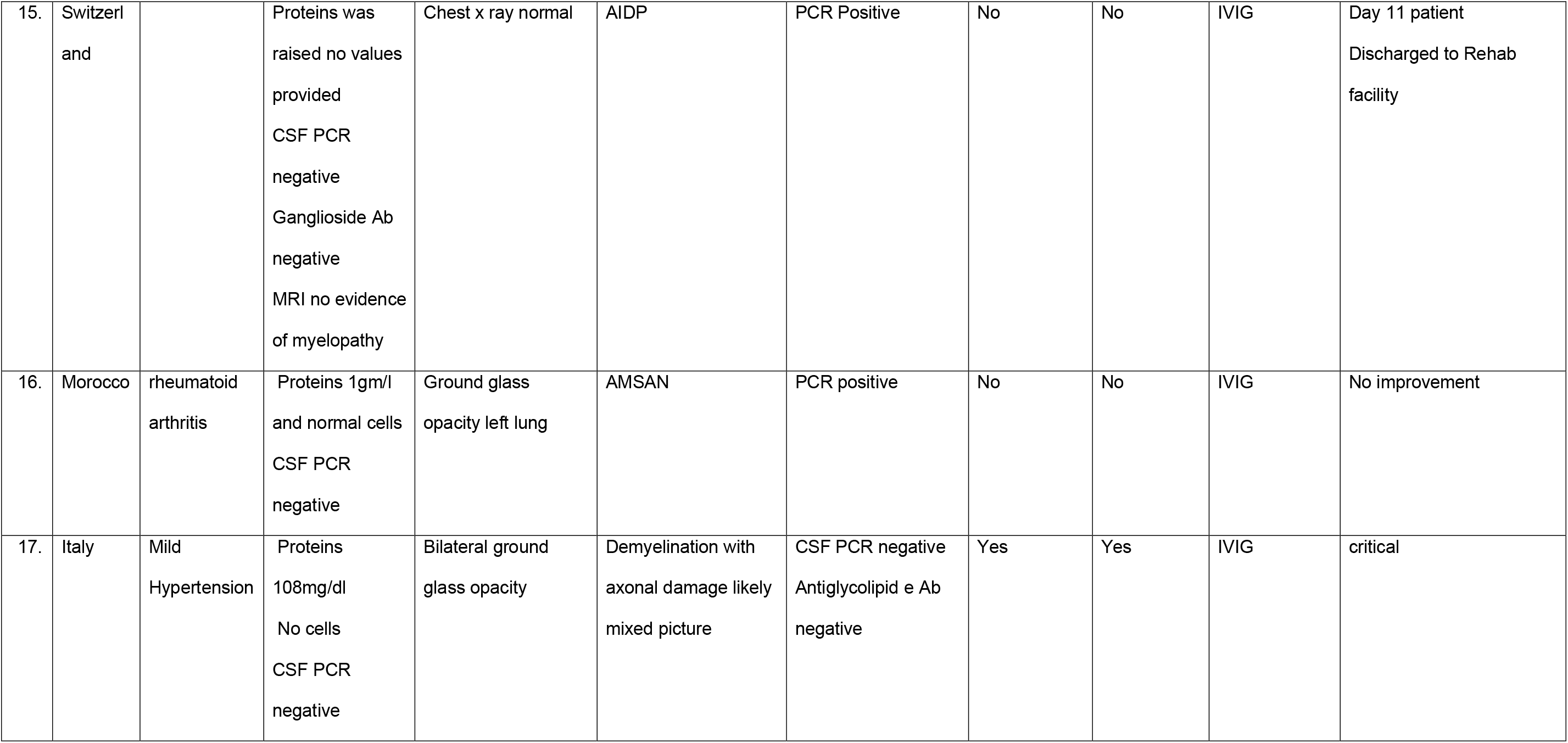
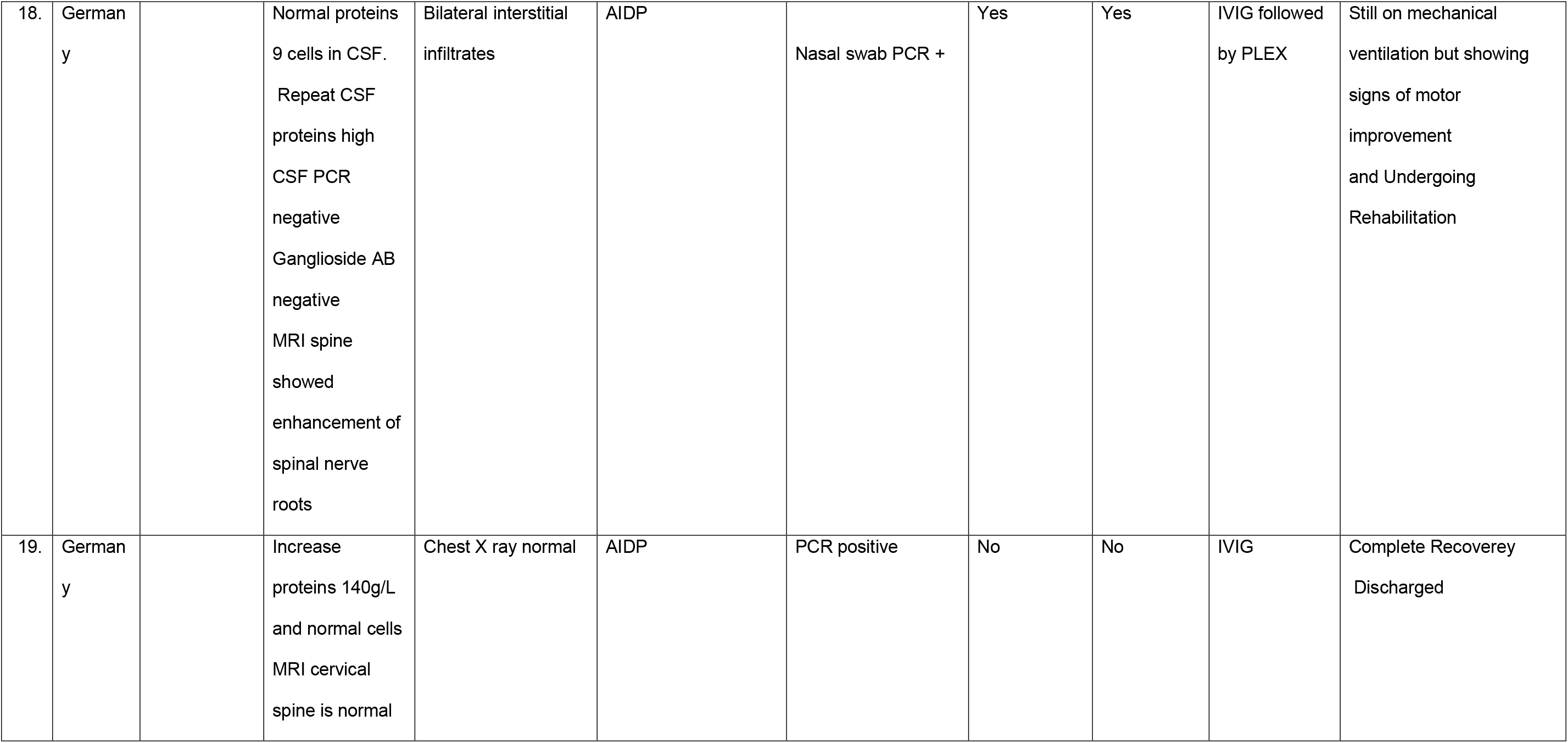
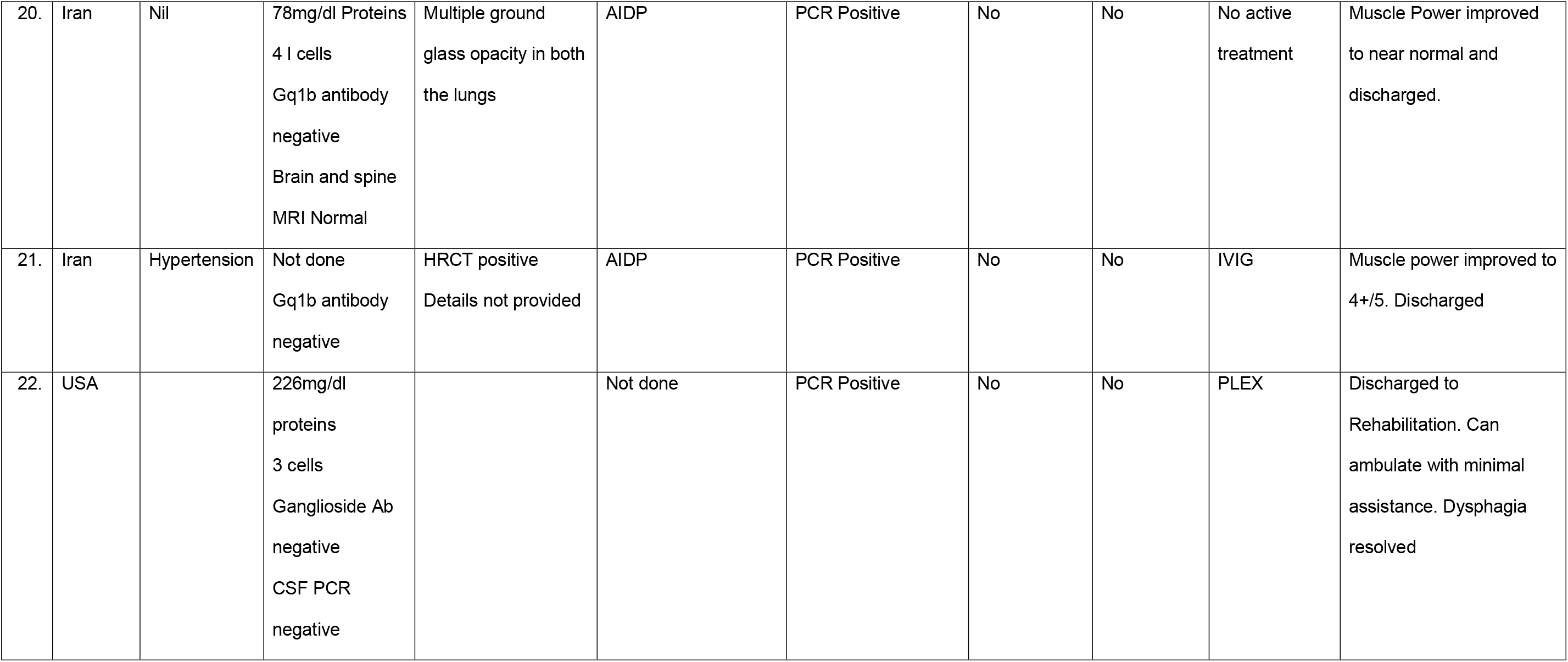
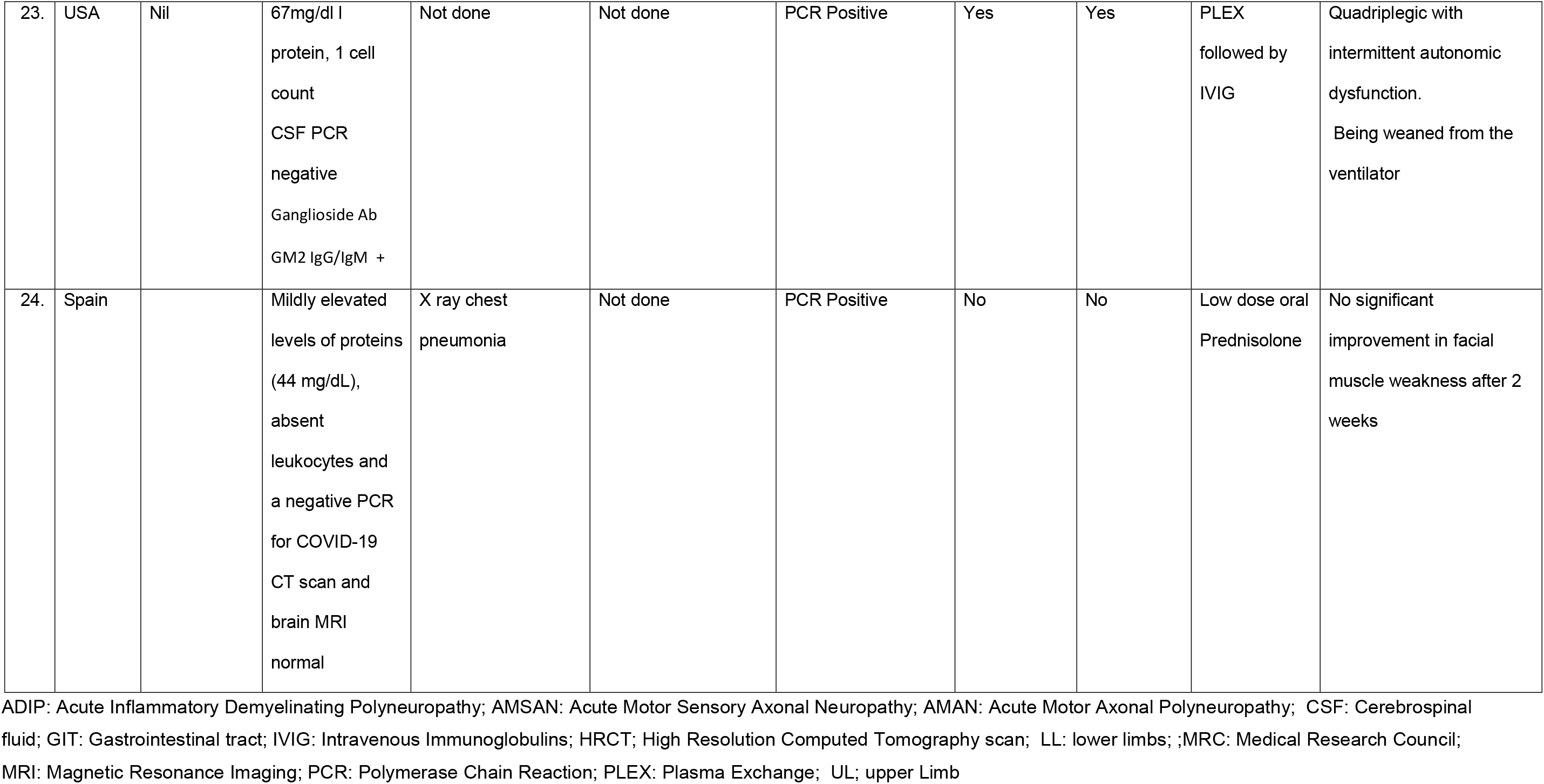
Details of the diagnostics, management, and outcomes of COVID-19 related GBS

### Demographics

Most of the cases (8) were reported from Italy ^26,27,28,29^ followed by the USA (4 cases)^30,31, 32^ Iran (3 cases),^33, 34^ Spain (3 cases),^35, 36^ Germany (2 cases) ^37,38^ and one case each from China ^39^, France ^40^, Switzerland ^41^ and Morocco. ^42^ Majority of the patients were males (18 /75%). The age ranged from 23-84 years and mean age is 60 years.

### Clinical Features

Most of the patients (17/24) had typical presenting features of GBS with sensory paresthesia followed by ascending paralysis. Three patients had Miller Fisher variant presenting as ataxia, ophthalmoplegia and areflexia. Total of nine patient developed facial palsy out of which six had bilateral facial palsy. One case had only bilateral facial palsy without any peripheral manifestations and was labeled as facial diplegic variant of GBS. Two patients developed bilateral six nerve palsy. One patient among above who initially presented with bilateral facial and hypoglossal palsy and progressed to a locked in syndrome like condition. One case each from the US^31^ and Switzerland ^41^ initially presented with paraparesis and bladder and bowel dysfunction. Spinal cord imaging was normal in both these cases and these were labeled as paraparetic variant with autonomic dysfunction.

An important peripheral nervous manifestation i.e. hyposmia and hypogeusia was reported in Five patients. One of them had complete reversal of hyposmia at the time of discharge. The predominant clinical presentation in majority of the cases was post-infectious. However, in three cases the onset of symptoms suggested a para-infectious course of disease.

### Laboratory and Radiological Investigations

Nasopharyngeal swab samples of all cases were PCR positive for COVID-19, except one case. That patient repeatedly tested negative but, later his serology tested positive for COVID-19. CSF PCR for COVID-19 was tested in twelve patients and it was negative in all. Ganglioside antibody was tested in twelve cases. It was positive in two case only. One of them was the Miller Fisher Variant. CSF analysis performed in 20 cases. Four patients had a normal CSF analysis while in 16 cases it showed albuminocytological disproportion of GBS.

An important finding was that revealed COVID-19 associated lung changes on High-Resolution Chest Tomography (HRCT) chest in fourteen cases. X-ray chest was normal in six cases and revealed pneumonia in one case. Chest imaging was not reported in two cases. This can potentially guide the clinicians. During this pandemic, in a GBS patient HRCT chest should be ordered in case of any doubt to detect possible COVID-19 associated pneumonia as both can be contribute towards respiratory failure.

### Electrodiagnostic Findings (EDX)

Nerve conduction studies/ Electromyography (NCS/EMG) were performed in 17 cases. Out of these twelve cases had prolong distal latencies (DML) and absent F waves suggestive of a typical demyelinating polyneuropathy. Four cases had Acute Motor and Sensory Axonal Neuropathy (AMSAN) variant and one had Acute Motor Axonal Neuropathy (AMAN) variant of GBS. However, in the Italian case series, the author reported their NCS/EMG as a mixed picture whereas a prolong DML and absent F waves favors a demyelinating variant.^28^

### Treatment

Three cases were ambulatory with minimum motor deficit and were not offered any treatment for GBS. One of them was a Miller Fisher variant. Nineteen patients were given IVIG. Among these two cases had repeat sessions of IVIG and two cases had PLEX after IVIG due to initial inadequate response. Two case had PLEX sessions as primary treatment one among them had IVIG after PLEX also.

### Outcomes and discharge status

One case expired due to complications. Nine patients were either discharged to nursing homes or shifted to rehabilitation for exercise. Complete recovery was reported in eight patients. At the time of publication of cases, three patients were on mechanical ventilation, one was critically ill, and no improvement was reported in one case. Outcome and discharge status were not mentioned for three cases.

## Discussion

This review suggests that COVID-19 associated GBS has emerged as an important neurological manifestation and complication of this global pandemic. Although, a clear association of COVID-19 leading to triggering of GBS is lacking at present, past experience with Zika Virus associated GBS suggests a possibility of causality between GBS and COVID-19 infection.

The onset of GBS was post infectious in all the cases in this review, except three in which it was para infectious. A similar pattern has also been seen in Zika Virus infections. Therefore, the treating physician should have a high index of suspicion in managing such cases. The patient might be in the infective stage of COVID-19 and proper barrier precautions will be necessary for the safety of hospital staff. Lung changes due to COVID-19 infection were see in a large number of patients in this review (15/24). Fourteen had a positive HRCT and one had pneumonia on X ray chest. All of these cases had a positive Nasopharyngeal PCR. Therefore, it is important to consider that during the current pandemic respiratory compromise in GBS may not be entirely due to neuromuscular failure but may also be due to COVID-19 pneumonia. At the same time if the patient with COVID-19 is having deterioration of respiratory function or is difficult to wean from ventilator GBS should also be considered as one of the possible reason.

This review suggests that in COVID-19 associated GBS, AIDP variant is more common followed by AMAN and AMSAN variants. However, Umapathi has recently suggested that Covid-19 GBS might be axonal due to paranodal pathologies and follow up EDX might help in reaching to a conclusion.^43^ Three patients presented with Miller Fisher syndrome variant which is a large number considering the very low incidence of Miller Fisher syndrome variant of GBS in general population.

The experience with Zika virus related GBS suggests that the patient present with typical symptoms including facial palsy on presentation, male predominance, and AIDP on EDX. A similar pattern was documented in this review. In a review from Puerto Rico facial weakness was seen in 62% cases of Zika Virus associated GBS as compared with 10% in non-Zika related GBS^44^. In this review 37.5 % of the cases had facial weakness with 5 having bilateral facial paralysis. The incidence of dysphagia in Zika Virus associated GBS has been reported to be 53.5% while it is very low in COVID-19 associated GBS (5/24 (20%).

Two patients had paraparesis at presentation followed by urinary retention and were later diagnoses as GBS. ^31,41^ This paraparetic pattern is seen more commonly in Zika Virus associated GBS cases. We do not know the exact mechanism of this phenomenon.

In 5 cases hyposmia and hypogeusia were either the presenting or co-existing features. ^29,35^ These were likely due to the COVID-19 infection and not because of GBS. This is an important finding and can be used as a clinical indicator of COVID-19 infections in suspected GBS cases.

Seropositivity of GBS for ganglioside antibody is reported to be around 30% with the cases of MFS having 95% GQ1b positivity. In this review only one case was positive GD 1b ganglioside antibody. However, this data is too small to make a conclusion Most of the cases in this review were treated with 5 sessions of IVIG. In two cases, IVIG was repeated while in two cases PLEX was also done after giving IVIG. PLEX has been used in two cases as initially and in one it was followed by IVIG due to inadequate response. One of the possible reasons for use of frequent use of IVIG in all these cases is that all of them were in high income countries with adequate resources and easy access to IVIG. We would like to suggest that in resource constrained areas and the developing world PLEX might prove to be equally beneficial as this is the preferred mode of treatment in cytokine storm syndrome due to COVID-19.

Most of the patients had a good outcome and were either discharged to home with complete recovery or were referred to rehabilitation for management of residual weakness and motor deficits. There was one death and four patients were reported to be on mechanical ventilation at the time of publication of the case reports. However, due to the limited data, it is difficult to comment if COVID-19 associated GBS increases severity of illness, length of Intensive care Unit (ICU) admission and prolongs ventilatory support.

### Comparison of MERS associated GBs Vs. and COVID-19 associated GBS

The published data regarding neurological complication and manifestations associated with MERS is limited. ^45,^**Error! Bookmark not defined**.^,47^ In addition, MERS was an epidemic limited to one geographic area and GBS associated with MERS was rarely reported so it is not possible to make a detailed comparison between this and COVID-19 associated GBS due to paucity of data. There is one case report of a critical illness neuropathy due to prolong intensive care unit stay reported from Saudi Arabia. ^46^ Kim *et.al* identified only four cases from Korea during the 2015 our break of MERS, which presented with neurological features. ^45^ One was diagnosed as GBS Bickerstaff variant, second one as Intensive care unit associated neuropathy overlapping with GBS and last 2 were labeled as toxic neuropathy. All four had sensory features on presentation and one of them developed motor weakness and ophthalmoplegia. However, EDX evaluation and CSF examination was normal in all patients. Ganglioside antibody was also negative. Only one patient required mechanical ventilation and was given IVIG. The other three did not have motor weakness and were only kept under medical observation, provided supplemental oxygen and no specific treatment was offered. These epidemics limited to a specific geographic zone affecting only 2494 people (WHO estimates), unlike COVID-19 which is a global health care crisis affecting millions. However, the common feature among both is the craniobulbar involvement in both. ^47^ See Table 3

**Table 3:** comparison of COVID-19 associated GBS with Zika associated GBS

| S No | Parameters | Zika Associated GBS – Puerto Rico <sup>44</sup> | Zika Associated GBS <sup>48</sup> | COVID-19 associated GBS |
| --- | --- | --- | --- | --- |
| 1. | Number of reported cases | 71 | 23 | 24 |
| 2. | Mean Age | 54 Years | 61 Years | 60 Years |
| 3. | Gender Distribution females | 37(52.1) | 8(34) | 6(25%) |
| 4. | Facial weakness | 44(62%) | 9(39%) | 9(37%) |
| 5. | Dysphagia | 38(53%) | 16(70%) | 2(8%) |
| 6. | Cough | 3(4.2%) |  | 21(87%) |
| 7. | Shortness of breath | 33(46.5%) | Not mentioned |  |
| 8. | Hyporeflexia/areflexia | 71(100) | 23(100%) | 23(99%) |
| 9. | Antecedent illness to neurological signs | 7 (0-21 ) | 5.9 (1.5–6.5) | 11(3-28) |
| 10. | Antecedent illness fever | 28(39.4) | 5(22%) | 18(72%) |
| 11. | Elevated proteins in CSF | 49/52(94.2%) | Not reported | 16(80%)<br>Not done in 4<br>Normal in 4 |
| 12. | ICU admission | 47(66.2%) | 14(61%) | 9(36%) |
| 13. | Mechanical ventilation | 22(31%) | 10(43%) | 9(36%) |
| 14. | Outcomes and discharge to rehab/nursing home | 35 (49.3) | Not mentioned | 3 still critical<br>2 no improvement<br>3 not reported |
| 15. | Discharge home | 32 (45.1) | Not mentioned | 16 |
| 16. | Death | 2 (2.8%) | 2 | 1 |

### Comparison of Zika Virus associated GBS Vs. COVID-19 associated GBS

The comparison of COVID-19 associated GBS with Zika Virus associated is presented in Table 3. In Zika Virus-GBS the median time from symptoms to disease onset was seven days consistent with para infectious GBS whereas in this review the median time was 11 days (3-28) days. In Zika Virus GBS the disease was more aggressive with frequent ICU admission and need for ventilatory support. Our data reports a similar pattern with a total of nine patients needing respiratory support. Seven were placed on mechanical ventilation and two were on noninvasive ventilation. On EDX evaluation demyelinating type is the most finding both with =Zika Virus and COVID-19 associated GBS. Cranial involvement is another feature common to both types of GBS.

### Limitations

Despite a rigorous search methodology for this review, we were not able to perform literature search all the major English bio-medical literature search databases due to lack of resources and access. There is a possibility that we might have missed some cases which we hope will be identified in the systematic reviews registered in the International prospective register of systematic reviews. The total number of confirmed cases of COVID-19 globally is now more than 7 million but we were able to document only 24 cases of GBS reported in the English biomedical literature so far. This is a very small number of cases to make a causal relationship or a definitive conclusion regarding COVID-19 associated GBS. Due to the wide spread of the disease and wide variations in the documentation and reporting of data from different parts of the world, there are chances that mild cases of GBS or cases with limited involvement might be missed or do not report to hospitals. Moreover, neurological services are not widely available in many developing countries and there is a possibility that some COVID-19 associated GBS cases remain undiagnosed due to lack of expertise in neurology. In addition, mortality in COVID-19 cases due to rapidly progressive respiratory failure is usually attributed to the COVID-19 itself. There is a possibility of co-existing GBS which may contribute to the worsening of the condition. We hope that as more data from different parts of the world is shared, things will become clearer in future and provide further insights into the COVID-19 associated GBS..

## Conclusion

The primary presentation of COVID-19 is respiratory but neurological manifestations and complications are increasingly being reported in the literature. GBS is one of the frequent neurological complication associated with COVID-19. There is no clear causative relationship between GBS, and COVID-19 at present and more data are needed to establish the casualty. However, from the available data we conclude that most of the cases present as a post-infectious disease with male preponderance. The EDX reveal a demyelinating type of polyneuropathy in most of the cases with few being AMAN and AMSAN variants. IVIG is the preferred mode of treatment and prognosis is generally good with most of the patients responding to treatment and rehabilitation plan. There is a need for large scale data collection on GBS and other related neurological manifestations and complications of COVID-19 to formulate better care plans in future.

## Data Availability

The scoping review is based on an online literature search. I have the full-text PDF of all articles cited in this manuscript

## Acknowledgements

Nil

## References

1. World Health Organization. Coronavirus disease (COVID-19) Situation Report – 142. Available from https://www.who.int/docs/default-source/coronaviruse/situation-reports/20200610-covid-19-sitrep-142.pdf?sfvrsn=180898cd_2 Accessed, June 11, 2020.

2. Mao L, Jin H, Wang M, et al. Neurologic Manifestations of Hospitalized Patients With Coronavirus Disease 2019 in Wuhan, China [published online ahead of print, 2020 Apr 10]. JAMA Neurol. 2020;e201127. doi:10.1001/jamaneurol.2020.1127

3. Helms J, Kremer S, Merdji H, Clere-Jehl R, Schenck M, Kummerlen C et.al.Neurologic Features in Severe SARS-CoV-2 Infection. N Engl J Med. 2020;382(23):2268–2270. doi:10.1056/NEJMc2008597.

4. Ahmad I, Rathore FA. Neurological manifestations and complications of COVID-19: A Literature Review.J Clin Neurosci 2020 May 6;S0967-5868(20)31078-X. doi:10.1016/j.jocn.2020.05.017. Online ahead of print.

5. Mathis S, Soulages A, Vallat JM, Le Masson G. History of acute polyradiculoneuropathy (part 1): The prehistory of Guillain-Barré syndrome. Neurology. 2020;94(19):828–35.

6. Mathis S, Soulages A, Le Masson G, Vallat JM. History of acute polyradiculoneuropathy (part 2): From 1916 to 2019. Neurology 2020;94(19):836–40.

7. Hughes RA, Cornblath DR. Guillain-barre syndrome. The Lancet. 2005;366(9497):1653–66.

8. Hughes RA, Rees JH. Clinical and epidemiologic features of Guillain-Barré syndrome. J Infect Dis. 1997;176 Suppl 2:S92–S98. doi:10.1086/513793

9. Yuki N, Hartung HP. Guillain–Barré syndrome. N Engl J Med. 2012;366(24):2294–304.

10. van den Berg B, Walgaard C, Drenthen J, Fokke C, Jacobs BC, van Doorn PA. Guillain-Barré syndrome: pathogenesis, diagnosis, treatment and prognosis. Nat Rev Neurol. 2014;10(8):469–482. doi:10.1038/nrneurol.2014.121

11. Ansari B, Basiri K, Derakhshan Y, Kadkhodaei F, Okhovat AA. Epidemiology and Clinical Features of Guillain-Barre Syndrome in Isfahan, Iran. Adv Biomed Res. 2018;7:87. Published 2018 May 29. doi:10.4103/abr.abr_50_17

12. Willison HJ, Jacobs BC, Van Doorn PA. Guillain-barre syndrome. The Lancet. 2016;388(10045):717–27.

13. Khan M, Muhammad W, Nawaz K, Ahmad I, Yousaf, M, Ahmad N, Rathore F, Ahmad K. FREQUENCY OF AXONAL VARIANTS OF GUILLAIN-BARRÉ SYNDROME IN PAKISTAN. PAFMJ [Internet]. 30 Sep.2011 [cited 11Jun.2020];61(3). Available from: https://www.pafmj.org/index.php/PAFMJ/article/view/804

14. Wu X, Wu W, Wang Z, et al. More severe manifestations and poorer short-term prognosis of ganglioside-associated Guillain-Barré syndrome in Northeast China. PLoS One. 2014;9(8):e104074. Published 2014 Aug 1. doi:10.1371/journal.pone.0104074

15. Zhang G, Li Q, Zhang R, Wei X, Wang J, Qin X. Subtypes and Prognosis of Guillain-Barré Syndrome in Southwest China. PLoS One. 2015;10(7):e0133520. Published 2015 Jul 22. doi:10.1371/journal.pone.0133520

16. Ashour HM, Elkhatib WF, Rahman MM, Elshabrawy HA. Insights into the Recent 2019 Novel Coronavirus (SARS-CoV-2) in Light of Past Human Coronavirus Outbreaks. Pathogens. 2020;9(3):186. Published 2020 Mar 4. doi:10.3390/pathogens9030186

17. Cui J, Li F, Shi ZL. Origin and evolution of pathogenic coronaviruses. Nat Rev Microbiol. 2019;17(3):181–192. doi:10.1038/s41579-018-0118-9

18. Baig AM, Khaleeq A, Ali U, Syeda H. Evidence of the COVID-19 Virus Targeting the. CNS: Tissue Distribution, Host-Virus Interaction, and Proposed Neurotropic Mechanisms. ACS Chem Neurosci. 2020;11(7):995–998. doi:10.1021/acschemneuro.0c00122

19. Natoli S, Oliveira V, Calabresi P, Maia LF, Pisani A. Does SARS-Cov-2 invade the brain? Translational lessons from animal models [published online ahead of print, 2020 Apr 25]. Eur J Neurol. 2020;10.1111/ene.14277. doi:10.1111/ene.14277

20. Arabi YM, Harthi A, Hussein J, Bouchama A, Johani S, Hajeer AH, et.al. Severe neurologic syndrome associated with Middle East respiratory syndrome corona virus (MERS-CoV). Infection. 2015 Aug 1;43(4):495–501.

21. Ng Kee Kwong KC, Mehta PR, Shukla G, Mehta AR. COVID-19, SARS and MERS: A neurological perspective [published online ahead of print, 2020 May 5]. J Clin Neurosci. 2020;S0967-5868(20)31185-1. doi:10.1016/j.jocn.2020.04.124

22. Finsterer J, Scorza FA, Ghosh R. COVID-19 polyradiculitis in 24 patients without SARS-CoV-2 in the cerebro-spinal fluid [published online ahead of print, 2020 Jun 4]. J Med Virol. 2020;10.1002/jmv.26121. doi:10.1002/jmv.26121

23. Ye Q, Wang B, Mao J. The pathogenesis and treatment of the ‘Cytokine Storm’ in COVID-19. J Infect. 2020;80(6):607–613. doi: 10.1016/j.jinf.2020.03.037. Epub 2020 Apr 10. PMID: 32283152; PMCID: PMC7194613.

24. Abdullahi A, Candan S, Elbol N, Olumide D. Guillian Barre syndrome in patients with COVID-19: a systematic review. PROSPERO 2020 CRD42020184822 Available from: https://www.crd.york.ac.uk/prospero/display_record.php?ID=CRD42020184822

25. Carrillo-Larco RM, Altez-Rodriguez C. COVID-19 and Guillain-Barre syndome: a systematic review of case reports. PROSPERO 2020 CRD42020182015 Available from: https://www.crd.york.ac.uk/prospero/display_record.php?ID=CRD42020182015

26. Padroni M, Mastrangelo V, Asioli GM, et al. Guillain-Barré syndrome following COVID-19: new infection, old complication? [published online ahead of print, 2020 Apr 24]. J Neurol. 2020;1–3. doi:10.1007/s00415-020-09849-6

27. Alberti P, Beretta S, Piatti M, et al. Guillain-Barré syndrome related to COVID-19 infection. Neurol Neuroimmunol Neuroinflamm. 2020;7(4):e741. Published 2020 Apr 29. doi:10.1212/NXI.0000000000000741

28. Ottaviani D, Boso F, Tranquillini E, Gapeni I, Pedrotti G, Cozzio S, et al. Early Guillain-Barré syndrome in coronavirus disease 2019 (COVID-19): a case report from an Italian COVID-hospital. Neurol Sci. 2020 Jun;41(6):1351–1354. doi:10.1007/s10072-020-04449-8..

29. Toscano G, Palmerini F, Ravaglia S, Ruiz L, Invernizzi P, Cuzzoni MG,, et al. Guillain-Barré Syndrome Associated with SARS-CoV-2 N Engl J Med. 2020;NEJMc2009191. doi:10.1056/NEJMc2009191. Epub ahead of print

30. Dinkin M, Gao V, Kahan J, Bobker S, Simonetto M, Wechsler P, Harpe J, Greer C et.al. COVID-19 presenting with ophthalmoparesis from cranial nerve palsy. Neurology. 2020 May 1:10.1212/WNL.0000000000009700. doi:10.1212/WNL.0000000000009700. Epub ahead of print.

31. Virani A, Rabold E, Hanson T, Haag A, Elrufay R, Cheema T, et al. Guillain-Barré Syndrome associated with SARS-CoV-2 infection [published online ahead of print, 2020 Apr 18]. IDCases. 2020;20:e00771. doi:10.1016/j.idcr.2020.e00771

32. Chan M, Han SC, Kelly S, Tamimi M, Giglio B, Lewis A. A case series of Guillain-Barré Syndrome following Covid-19 infection in New York. Neurology: Clinical Practice. 2020 May 20.

33. Sedaghat Z, Karimi N. Guillain Barre syndrome associated with COVID-19 infection: A case report. J Clin Neurosci. 2020;76:233–235. doi:10.1016/j.jocn.2020.04.062

34. Ebrahimzadeh SA, Ghoreishi A, Rahimian N. Guillain-Barré Syndrome associated with the coronavirus disease 2019 (COVID-19). Neurology: Clinical Practice. 2020 May 20.

35. Gutiérrez-Ortiz C, Méndez A, Rodrigo-Rey S, San Pedro-Murillo E, Bermejo-Guerrero L, Gordo-Mañas R,, et al. Miller Fisher Syndrome and polyneuritis cranialis in COVID-19 [published online ahead of print, 2020 Apr 17. Neurology. 2020;10.1212/WNL.0000000000009619. doi:10.1212/WNL.0000000000009619

36. Juliao Caamaño DS, Alonso Beato R. Facial diplegia, a possible atypical variant of Guillain-Barré Syndrome as a rare neurological complication of SARS-CoV-2. J Clin Neurosci. 2020 May 14. doi:10.1016/j.jocn.2020.05.016. Epub ahead of print.

37. Scheidl E, Canseco DD, Hadji-Naumov A, Bereznai B. Guillain-Barré syndrome during SARS-CoV-2 pandemic: A case report and review of recent literature [published online ahead of print, 2020 May 10]. J Peripher Nerv Syst. 2020;10.1111/jns.12382. doi:10.1111/jns.12382

38. Pfefferkorn T, Dabitz R, von Wernitz-Keibel T, Aufenanger J, Nowak-Machen M, Janssen H. Acute polyradiculoneuritis with locked-in syndrome in a patient with Covid-19 [published online ahead of print, 2020 May 12]. J Neurol. 2020;1–2. doi:10.1007/s00415-020-09897-y

39. Zhao H, Shen D, Zhou H, Liu J, Chen S. Guillain-Barré syndrome associated with SARS-CoV-2 infection: causality or coincidence?. Lancet Neurol. 2020;19(5):383–384. doi:10.1016/S1474-4422(20)30109-5

40. Camdessanche J-Philippe, Morel J, Pozzetto B, Paul S, Tholance Y, Botelho-Nevers E, COVID-19 may induce Guillain-Barr ’e syndrome, Revue Neurologique (2020), doi: https://doi.org/10.1016/j.neurol.2020.04.003

41. Coen M, Jeanson G, Culebras Almeida LA, Hübers A, Stierlin F et.al. Guillain-Barré syndrome as a complication of SARS-CoV-2 infection. Brain Behav Immun. 2020:S0889-1591(20)30698-X. doi:10.1016/j.bbi.2020.04.074. Epub ahead of print.

42. Otmani HE, Moutawakil BE, Rafai M-Abdoh, Benna NE, Kettani CE, Soussi M, Mdaghri NE, Barrou H, Afif H, Covid-19 and Guillain-Barr ’e syndrome: More than a coincidence!, Revue Neurologique (2020), doi: https://doi.org/10.1016/j.neurol.2020.04.007

43. Umapathi T. Does COVID-19 cause axonal GBS? J Clin Neurosci. 2020 May 23:S0967-5868(20)31285-6. doi:10.1016/j.jocn.2020.05.057. Epub ahead of print.

44. Dirlikov E, Major CG, Medina NA, Lugo-Robles R, Matos D, Muñoz-Jordan JL, et all. Clinical features of Guillain-Barré syndrome with vs without Zika virus infection, Puerto Rico, 2016. JAMA neurology. 2018 Sep 1;75(9):1089–97

45. Kim JE, Heo JH, Kim HO, Song SH, Park SS, Park TH, et al. Neurological complications during treatment of Middle East respiratory syndrome. Journal of Clinical Neurology. 2017 Jul 1;13(3):227–33.

46. Arabi YM, Harthi A, Hussein J, Bouchama A, Johani S, Hajeer AH, et al. Severe neurologic syndrome associated with Middle East respiratory syndrome corona virus (MERS-CoV). Infection 2015;43:495–501

47. Zegarra-Valdivia J, Vilca BN, Tairo T, Munive V, Lastarria C. Neurological component in coronaviruses induced disease: systematic review of SARS-COV, MERS-COV, and SARS-CoV-2.

48. Rozé B, Najioullah F, Fergé JL, Dorléans F, Apetse K, Barnay JL, Daudens-Vaysse E, Brouste Y, Césaire R, Fagour L, Valentino R. Guillain-Barré syndrome associated with Zika virus infection in Martinique in 2016: a prospective study. Clinical Infectious Diseases. 2017 Oct 16;65(9):1462–8.

